# Development of a Novel Score to Predict Urgent Mechanical Circulatory Support in Chronic Total Occlusion Percutaneous Coronary Intervention

**DOI:** 10.1101/2023.02.03.23285426

**Authors:** Judit Karacsonyi, Larissa Stanberry, Bahadir Simsek, Spyridon Kostantinis, Salman S. Allana, Athanasios Rempakos, Brynn Okeson, Khaldoon Alaswad, Mir B. Basir, Farouc Jaffer, Paul Poommipanit, Jaikirshan Khatri, Mitul Patel, Ehtisham Mahmud, Abdul Sheikh, Jason R. Wollmuth, Robert W Yeh, Raj H. Chandwaney, Ahmed M ElGuindy, Nidal AbiRafeh, Daniel R. Schimmel, Keith Benzuly, M. Nicholas Burke, Bavana V. Rangan, Olga C. Mastrodemos, Yader Sandoval, Imre Ungi, Emmanouil S. Brilakis

## Abstract

**Background:** Estimating the likelihood of urgent mechanical circulatory support (MCS) can facilitate procedural planning and clinical decision making in chronic total occlusion (CTO) percutaneous coronary intervention (PCI).

**Methods:** We analyzed 2,784 CTO PCIs performed between 2012 and 2021 at 12 centers. The variable importance was estimated by a bootstrap applying a random forest algorithm to a propensity-matched sample (a ratio of 1:5 matching cases with controls on center). The identified variables were used to predict the risk of urgent MCS. The performance of the risk model was assessed in-sample as well as on 2411 out-of-sample procedures who did not require urgent MCS.

**Results:** Urgent MCS was used in 62 (2.2%) of cases. Patients who required urgent MCS were older (70 [63, 77] vs. 66 [58, 73] years, p=0.003) compared with those who did not require urgent MCS. Technical (68% vs. 87%, p<0.001) and procedural successes (40% vs. 85%, p<0.001) were lower in the urgent MCS group compared with no urgent MCS cases. The risk model for urgent MCS use included retrograde crossing strategy, left ventricular ejection fraction, and lesion length. The resulting model demonstrated good calibration and discriminatory capacity with AUC (95%CI) of 0.79 (0.73, 0.86) and specificity and sensitivity of 86% and 52%, respectively. On the out-of-sample set, the specificity of the model was 87%.

**Conclusion:** The PROGRESS CTO MCS score can help estimate the risk of urgent MCS use during CTO PCI.

**What Is Known?:**

- Estimating the likelihood of urgent mechanical circulatory support (MCS) can facilitate procedural planning and clinical decision making in chronic total occlusion (CTO) percutaneous coronary intervention (PCI).

**What the Study Adds?:**

- We developed a risk model for urgent MCS use during CTO PCI using retrograde crossing strategy, left ventricular ejection fraction, and lesion length.
- Use of the PROGRESS CTO urgent MCS score may facilitate patient selection for prophylactic hemodynamic support optimizing the risk benefit ratio of the procedure.

## INTRODUCTION

Chronic total occlusion (CTO) percutaneous coronary interventions (PCI) can be complex procedures with approximately 85-90% technical success rates at experienced centers but also relatively high incidence of major adverse cardiac events (1-3%).^1-4^ Urgent mechanical circulatory support (MCS) might be necessary in some complication cases. The use of mechanical circulatory devices to support high-risk elective PCI has become more common in part due to increasing number of patients considered inoperable or high risk for surgical revascularization.^5^ Estimating the need for urgent MCS could facilitate clinical decision-making and procedural planning in CTO PCI. We developed a score to identify patients at increased risk for urgent MCS.

## MATERIALS AND METHODS

We analyzed 2,784 CTO PCIs performed between 2012 and 2021 at 12 centers in the PROGRESS-CTO Registry (Prospective Global Registry for the Study of Chronic Total Occlusion Intervention; Clinicaltrials.gov identifier: NCT02061436); only data from centers with at least 40 PCIs and those with urgent MCS were used. Study data were collected and managed using REDCap (Research Electronic Data Capture) electronic data capture tools hosted at the Minneapolis Heart Institute Foundation.^6,7^ The study was approved by the institutional review board of each site.

Coronary CTOs were defined as coronary lesions with Thrombolysis in Myocardial Infarction (TIMI) grade 0 flow of at least 3-month duration. Estimation of the duration of occlusion was clinical, based on the first onset of angina, prior history of myocardial infarction (MI) in the target vessel territory, or comparison with a prior angiogram. Calcification was assessed by angiography as mild (spots), moderate (involving ≤50% of the reference lesion diameter), or severe (involving >50% of the reference lesion diameter). Moderate proximal vessel tortuosity was defined as the presence of at least 2 bends >70° or 1 bend >90° and severe tortuosity as 2 bends >90° or 1 bend >120° in the CTO vessel. A retrograde procedure was an attempt to cross the lesion through a collateral vessel or bypass graft supplying the target vessel distal to the lesion; otherwise, the intervention was classified as an antegrade-only procedure. Antegrade dissection/re-entry was defined as antegrade PCI during which a guidewire was intentionally introduced into the extraplaque space proximal to the lesion, or re-entry into the distal true lumen was attempted after intentional or inadvertent extraplaque guidewire crossing. Technical success was defined as successful CTO revascularization with achievement of <30% residual diameter stenosis within the treated segment and restoration of TIMI grade 3 antegrade flow.^8^ Procedural success was defined as achievement of technical success without any in-hospital major adverse cardiac event (MACE), that were defined as any of the following events prior to hospital discharge: death, MI, recurrent symptoms requiring urgent repeat target-vessel revascularization (TVR) with PCI or coronary artery bypass graft (CABG) surgery, tamponade requiring either pericardiocentesis or surgery, and stroke. MI was defined using the Third Universal Definition of Myocardial Infarction (type 4a MI).^9^ The PROGRESS-CTO score as described by Christopoulos et al ^10^ and PROGRESS MACE score was described by Simsek et al ^11^. Urgent mechanical circulatory support was defined as use of MCS after the procedure started and not in a planned fashion.

### Statistical Analysis

Categorical variables were expressed as counts (%) and compared using the Pearson’s chi-squared test or the Fisher’s exact test, as appropriate. Continuous variables in the study were skewed and were summarized by medians (interquartile ranges [IQR]); the variables were compared between urgent MCS and non-urgent MCS patients using the Wilcoxon rank-sum test.

To develop a prediction model to identify procedures that may require urgent MCS, first, risk factors were selected, then three competing models were built and their performances were compared. Given the small number of urgent MCS cases and sample imbalance, we used a random forest (RF) algorithm with a bootstrap (B=1000) data to identify risk factors to include in the model. For that, based on clinical reasoning and existing literature, the following factors plausibly associated with the risk of UCS were identified: age, gender, BMI, left ventricular ejection fraction (LVEF), creatinine levels at baseline (log scale), retrograde approach, prior MI, prior CABG, prior heart failure, prior PAD, CVD, diabetes mellitus, vessel diameter, proximal cap ambiguity, lesion length, side branch at proximal cap, and blunt stump. The importance of each risk factor was estimated by applying an RF algorithm to a propensity-matched bootstrap sample (n=373, with a ratio of 1:5 matching UCS with non-UCS patients on center only). The prediction error and the node impurity were estimated for these variables on each of the bootstrap samples using accuracy and the Gini index, accordingly. The variables were ranked by their index medians and the top five factors for each of the two importance measures were selected: retrograde approach, LVEF, proximal cap ambiguity, lesion length, and creatinine were the most important based on model accuracy and age, BMI, LVEF, creatinine, lesion length were the top for the Gini index of node impurities.

With risk factors identified, an independent matched sample (n=373) was drawn using the same criteria as above and two predictive models were built using a logistic regression approach for the two sets of variables; these models are referred to as M_1_ and M_2_ (for Gini and accuracy). A third joint model, M_3_, was built by applying a logistic regression with a backward elimination to a joint factor set across the first two models, i.e. retrograde approach, LVEF, proximal cap ambiguity, lesion length, creatinine, age, BMI. The three risk models were validated using a bootstrap resampling; the estimated shrinkage parameters and bias-corrected performance indices are reported. The indices were based on the receiver operating characteristic (ROC) curves including area under the curve (AUC), specificity, and sensitivity. The performance of the models was also assessed on the remaining sample of patients with no urgent MCS (n=2,411).

The statistical analysis was performed in R 4.1.2 (R Core Team) in RStudio 2022.07.1 environment (RStudio, PBC).

## RESULTS

Urgent MCS was used in 62 (2.2%) of 2,722 CTO PCIs. The baseline clinical characteristics of the study patients classified according to urgent MCS use and their angiographic characteristics are shown in Table 1. Urgent MCS patients were older (70 [63, 77] vs. 66 [58, 73] years, p=0.003) compared with patients who did not receive urgent MCS. Prior heart failure (34% vs. 28%, p=0.287) and prior myocardial infarction (53% vs. 41%, p=0.075) were similar between the two groups. Left ventricular ejection fraction was lower in the urgent MCS group (45% [33, 55] vs. 55% [43, 60], p<0.001). The CTO lesions in the urgent MCS group were more complex with higher prevalence of moderate/severe calcification (69% vs. 52%, p=0.007), moderate/severe tortuosity (47% vs. 31%, p=0.008) and proximal cap ambiguity (55% vs. 37%, p=0.006). Use of the retrograde approach (77% vs. 36%, p<0.001) was also more common in the urgent MCS group.

**Table 1.**
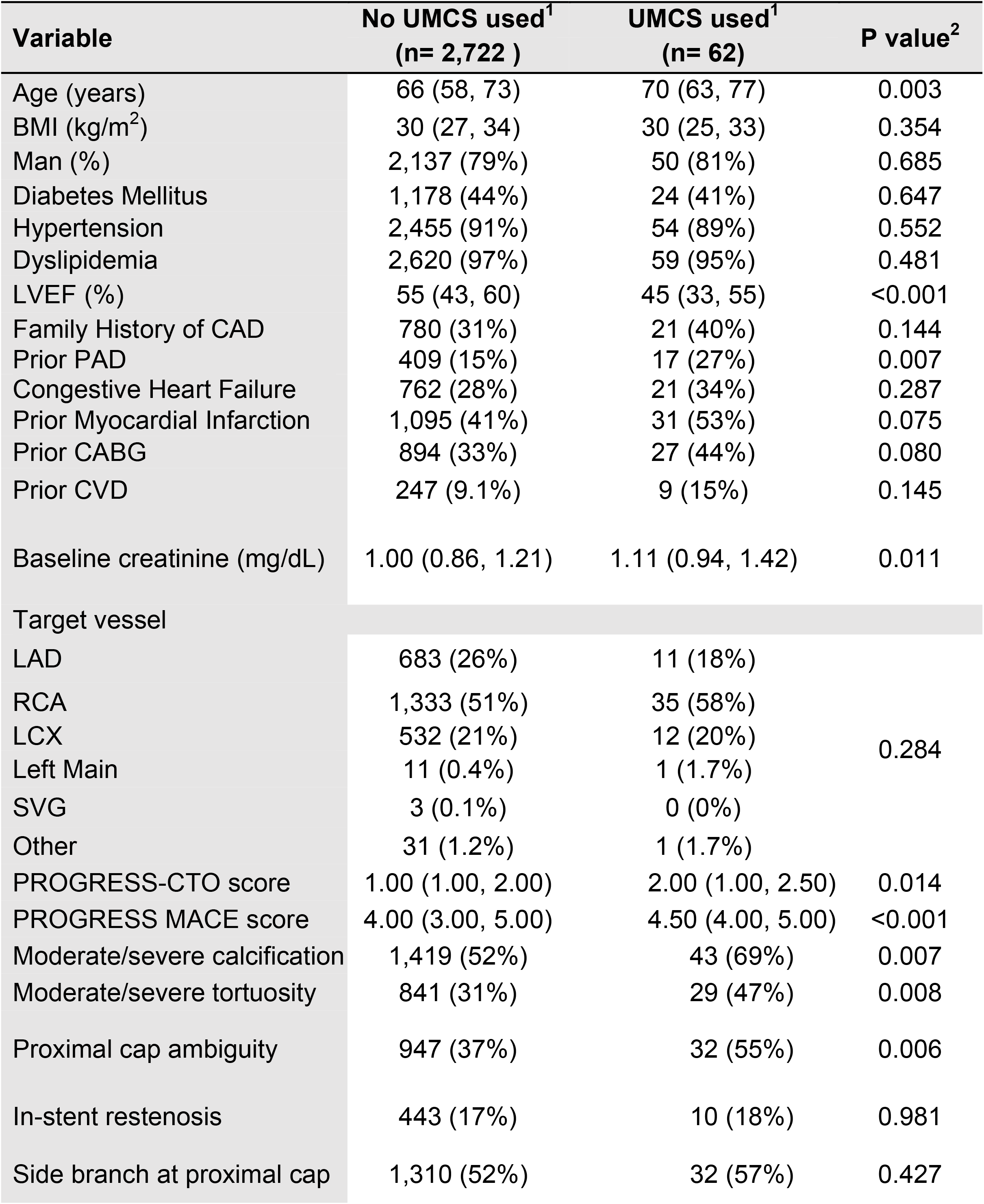
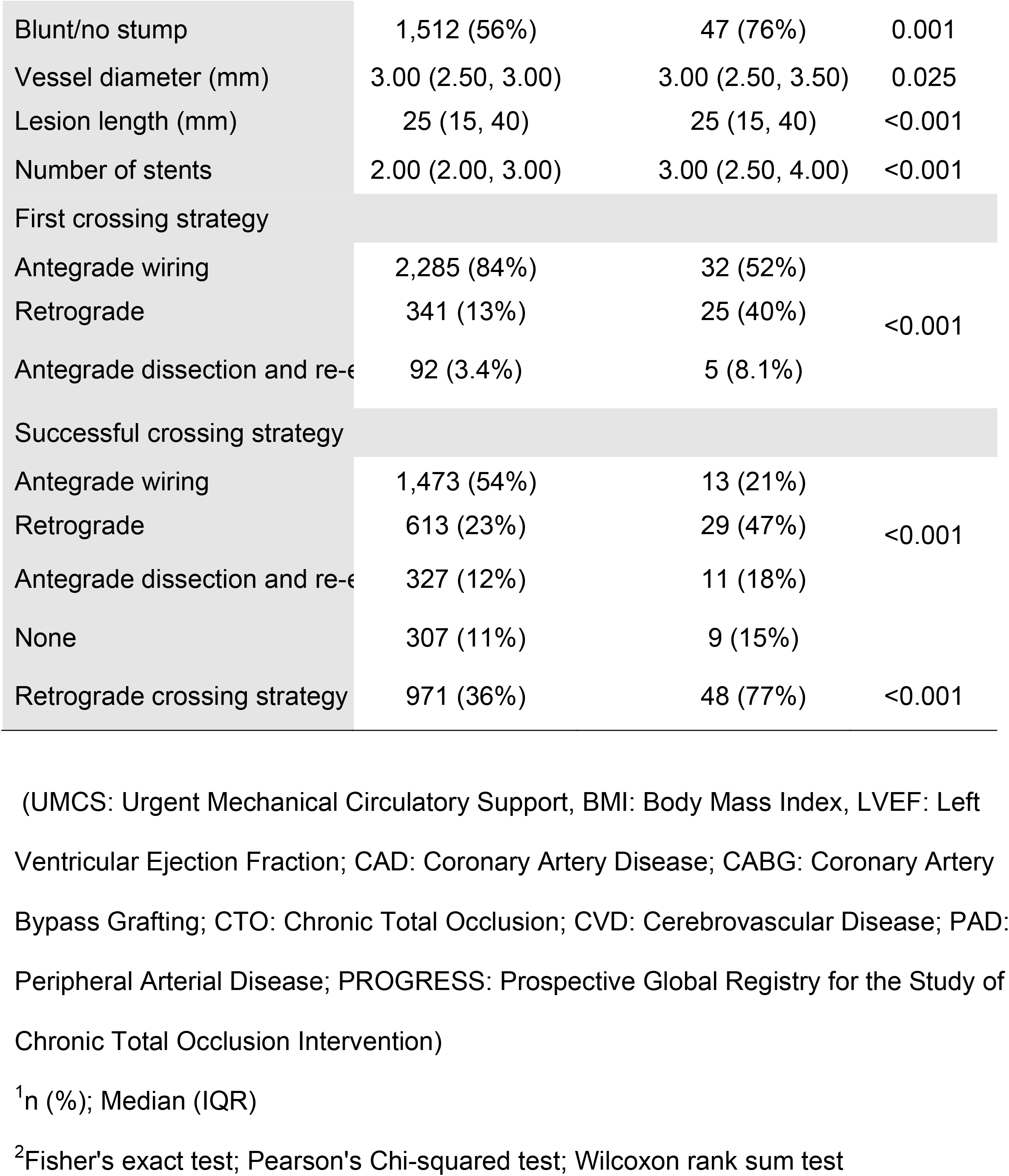
Baseline clinical characteristics of study patients with and without urgent mechanical support.

Procedural outcomes classified according to urgent MCS use are presented in Table 2. Technical (68% vs. 87%, p<0.001) and procedural success (40% vs. 85%, p<0.001) was lower and MACE was higher (31% vs. 2.1%, p<0.001) in the urgent MCS group. Urgent MCS was associated with longer procedural (256 [193, 328] min vs. 140 [98, 196] min, p<0.001) and fluoroscopy time 93 [60, 121] min vs. 51 [31, 79] min, p<0.001) and higher air kerma radiation dose 3.39 [2.09,4.81] Gray vs. 1.99 [1.14, 3.30] Gray, p<0.001) compared with no urgent MCS cases.

**Table 2.** Procedural characteristics and outcomes of study patients with and without urgent mechanical circulatory support (UMCS).

| Variable | No UMCS used<sup>1</sup><br>(n= 2,722 ) | UMCS used<sup>1</sup><br>(n= 62) | P value<sup>2</sup> |
| --- | --- | --- | --- |
| Technical Success | 2,373 (87%) | 42 (68%) | <0.001 |
| Procedural Success | 2,316 (85%) | 25 (40%) | <0.001 |
| Procedural time (min) | 140 (98, 196) | 256 (193, 328) | <0.001 |
| Fluoroscopy time (min) | 51 (31, 79) | 93 (60, 121) | <0.001 |
| Air kerma radiation dose (Gray) | 1.99 (1.14, 3.30) | 3.39 (2.09,4.81) | <0.001 |
| MACE | 57 (2.1%) | 19 (31%) | <0.001 |
| Death | 5 (0.2%) | 8 (13%) | <0.001 |
| Acute MI | 17 (0.6%) | 7 (11%) | <0.001 |
| Re-PCI | 7 (0.3%) | 2 (3.2%) | 0.016 |
| Stroke | 6 (0.2%) | 0 (0%) | >0.999 |
| Emergency CABG | 2 (<0.1%) | 1 (1.6%) | 0.065 |
| Pericardiocentesis | 28 (1.0%) | 7 (11%) | <0.001 |
| Perforation | 139 (5.1%) | 22 (35%) | <0.001 |
| Tamponade | 20 (0.7%) | 5 (8.1%) | <0.001 |
| Dissection/Thrombus of Donor Artery | 22 (0.8%) | 8 (13%) | <0.001 |
| Vascular access site complication | 41 (1.5%) | 4 (6.5%) | 0.017 |

(UMCS: Urgent Mechanical Circulatory Support; MACE: Major Adverse Cardiac Events;
MI: Myocardial Infarction; PCI: Percutaneous Coronary Intervention; CABG: Coronary Artery Bypass Grafting)
<sup>1</sup>n (%); Median (IQR)
<sup>2</sup>Pearson's Chi-squared test; Fisher's exact test, Wilcoxon rank sum test

The baseline clinical and angiographic characteristics of the study patients included in the prediction model are shown in Table 3. The outcomes of the procedures included in the model are demonstrated in Table 4.

**Table 3.**
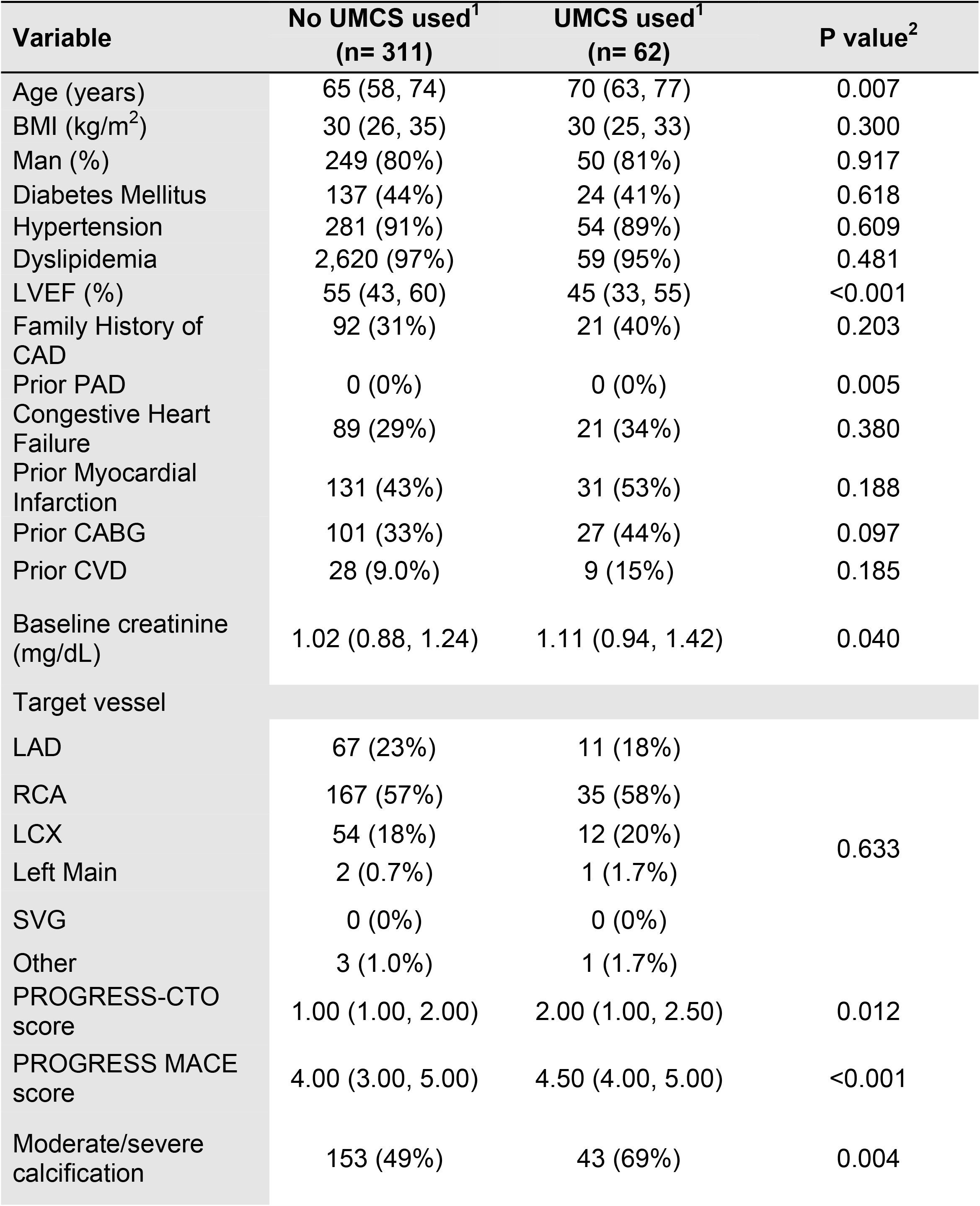
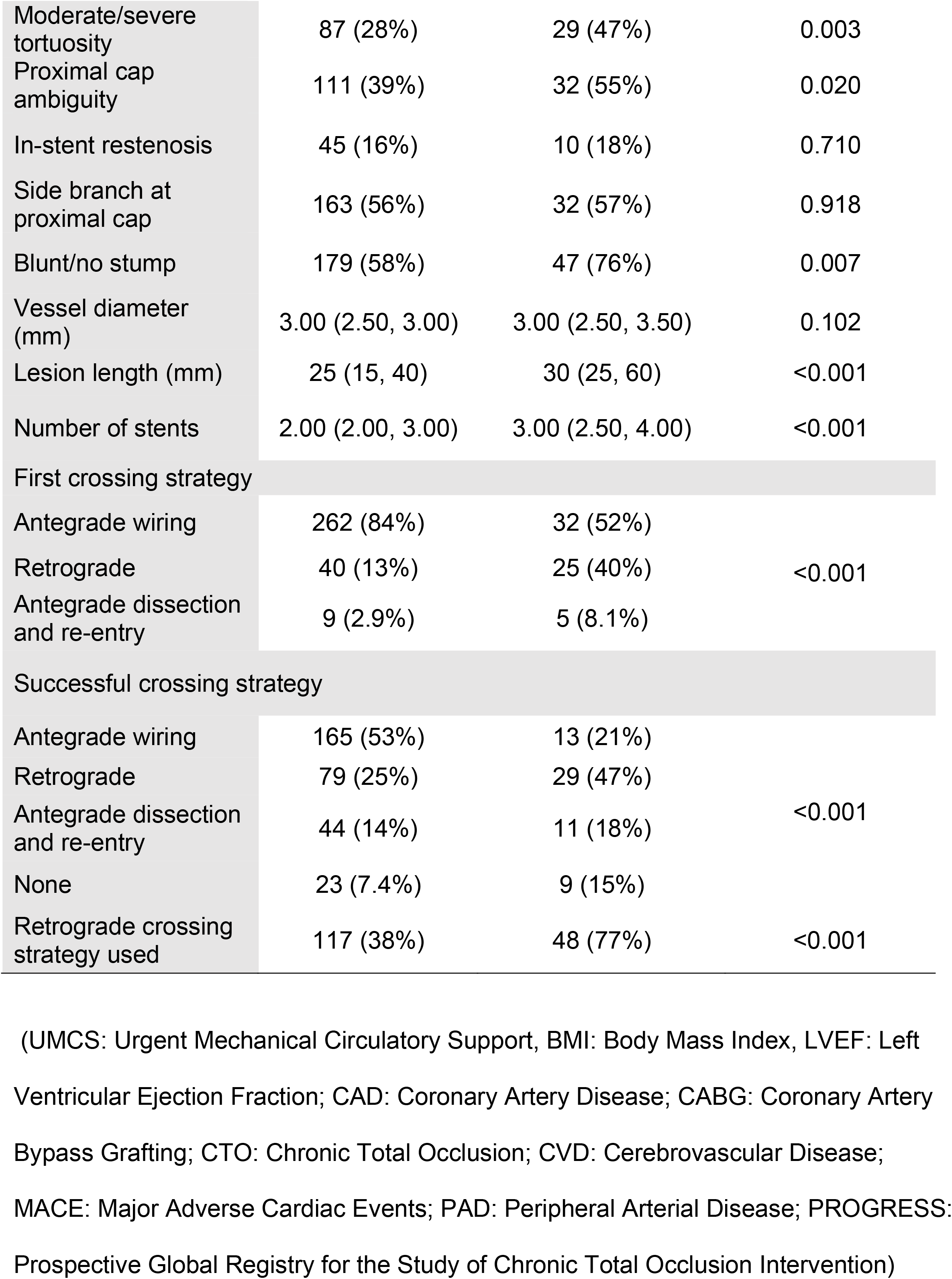
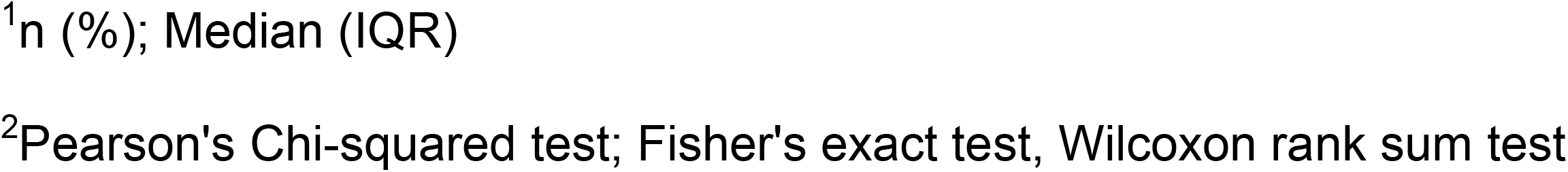
Baseline clinical and angiographic characteristics of study patients included in the model with and without urgent mechanical support.

**Table 4.** Procedural characteristics and outcomes of the cases included in the model with and without urgent mechanical support.

| Variable | No UMCS used<sup>1</sup><br>(n= 311 ) | UMCS used<sup>1</sup><br>(n= 62) | P value<sup>2</sup> |
| --- | --- | --- | --- |
| Technical Success | 285 (92%) | 42 (68%) | <0.001 |
| Procedural Success | 283 (91%) | 25 (40%) | <0.001 |
| Procedural time (min) | 140 (98, 196) | 256 (193, 328) | <0.001 |
| Fluoroscopy time (min) | 51 (31, 79) | 93 (60, 121) | <0.001 |
| Air kerma radiation dose (Gray) | 1.99 (1.14, 3.30) | 3.39 (2.09,4.81) | <0.001 |
| MACE | 4 (1.3%) | 19 (31%) | <0.001 |
| Death | 0 (0%) | 8 (13%) | <0.001 |
| Acute MI | 2 (0.6%) | 7 (11%) | <0.001 |
| Re-PCI | 0 (0%) | 2 (3.2%) | 0.027 |
| Stroke | 0 (0%) | 0 (0%) | >0.999 |
| Emergency CABG | 0 (0%) | 1 (1.6%) | 0.166 |
| Pericardiocentesis | 2 (0.6%) | 7 (11%) | <0.001 |
| Perforation | 16 (5.1%) | 22 (35%) | <0.001 |
| Tamponade | 2 (0.6%) | 5 (8.1%) | 0.002 |
| Dissection/Thrombus of Donor Artery | 2 (0.6%) | 8 (13%) | <0.001 |
| Vascular access site complication | 6 (1.9%) | 4 (6.5%) | 0.066 |

(UMCS: Urgent Mechanical Circulatory Support; MACE: Major Adverse Cardiac Events;
MI: Myocardial Infarction; PCI: Percutaneous Coronary Intervention; CABG: Coronary Artery Bypass Grafting)
†: median (interquartile ranges)
<sup>1</sup>n (%), Median (IQR)
<sup>2</sup>Pearson's Chi-squared test; Fisher's exact test, Wilcoxon rank sum test

The final joint model, M3, included retrograde approach, LVEF, and lesion length. The three models M1-M3 demonstrated reasonable calibration and discriminatory capacity (Figure 1). The estimated AUC (95%CI) for these models were 0.77 (0.70, 0.84), 0.78 (0.72, 0.85), and 0.79 (0.73, 0.86) with the shrinkage factors of 0.85, 0.90, and 0.93, respectively. Adjusting for optimism, the corrected AUC were 0.75, 0.76, and 0.79.

**Figure 1.**
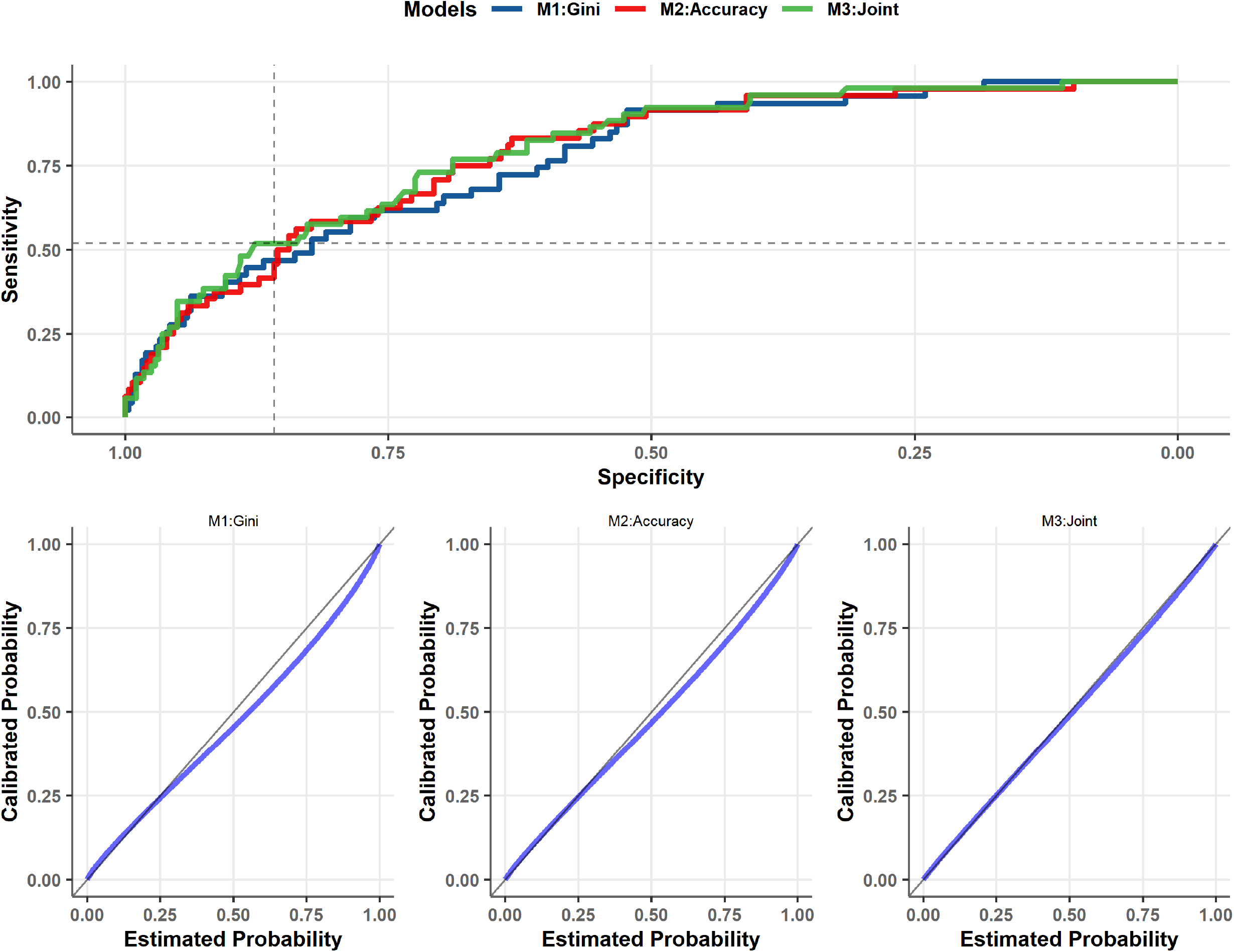
**Panel A:** Urgent mechanical circulatory support (MCS) use and receiver operator characteristics analyses of the PROGRESS CTO MCS score for models based on Gini index (M1), classification accuracy (M2), and using a joint set of M1/M2 variables (M3) **Panel B:** Calibration plots for M1-M3 models.

For the joint model, a threshold of 0.27 corresponded to accuracy (95%CI) of 80.6% (76.0, 84.6) with specificity of 85.8% and sensitivity of 51.9%. In the out-of-sample set of patients with no urgent MCS (n=2,411), the estimated specificity of the joint model was 86.8%, consistent with the in-sample validation. (The baseline characteristics and outcomes of the validation dataset are presented in Supplementary Tables 1.

Based on the joint model, a nomogram in Figure 2. gives a simple bedside tool to estimate the risk of urgent MCS.

**Figure 2.**
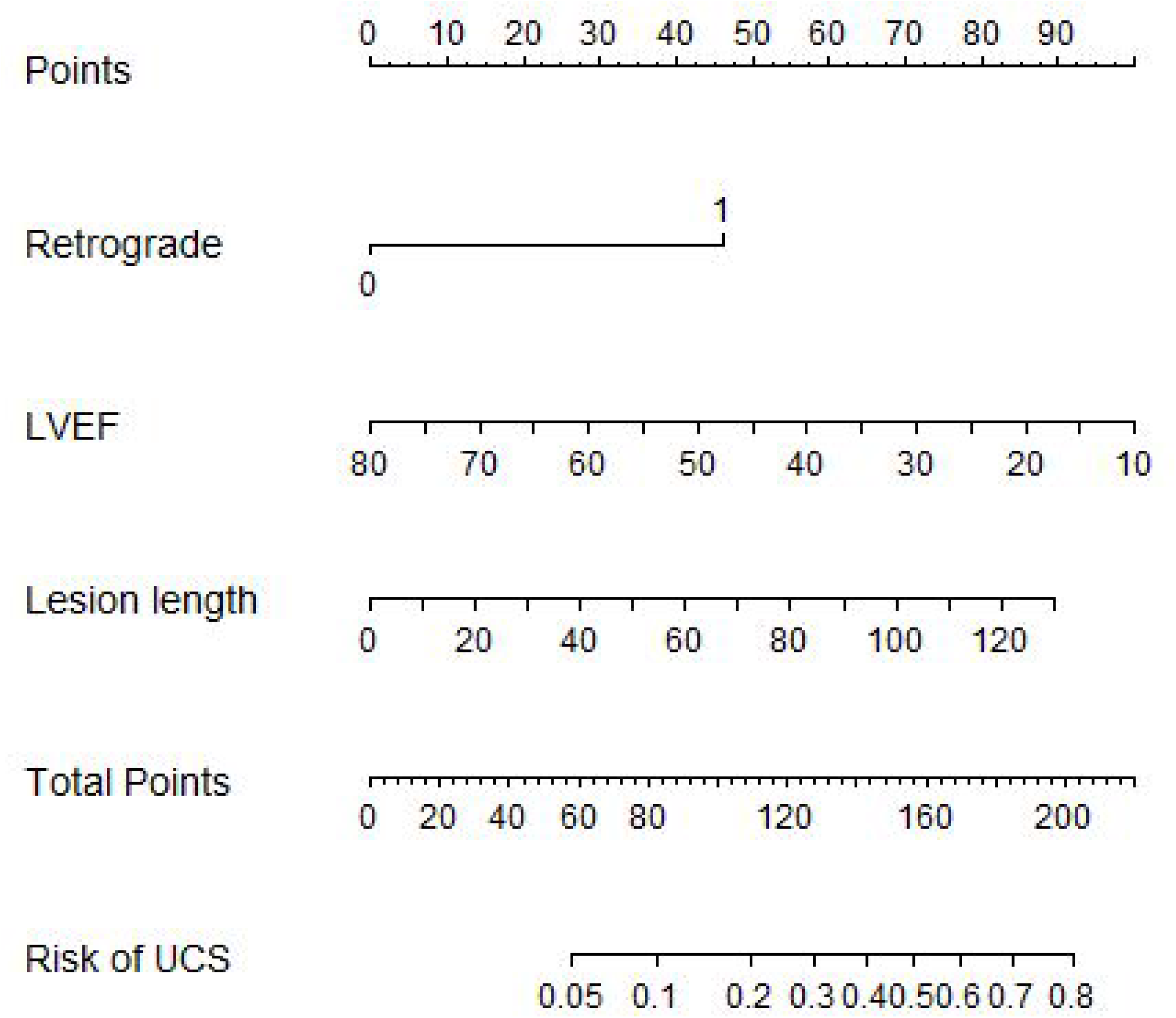
A nomogram of the PROGRESS CTO MCS score. Example patient: Use of retrograde crossing strategy (45 points), LVEF<30% (70 points), lesion length> 60 mm (40 points), this means a total of 155 points which translates to 0.5 (50%) risk of using urgent MCS.

## DISCUSSION

We developed a novel score for estimating the risk of urgent MCS during CTO PCI. The score may be a useful aid to assist in procedure planning.

The 2021 American College of Cardiology/American Heart Association/Society for Cardiovascular Angiography and Interventions Guideline for Coronary Artery Revascularization guidelines support elective insertion of appropriate mechanical circulatory support in selected high-risk patients, as an adjunct to PCI to prevent hemodynamic compromise during PCI with class IIB, level of evidence B recommendation.^12^ CTO PCI carries increased risk of complications compared with non-CTO PCI due to complex coronary anatomy (calcification, tortuosity, multivessel disease), difficulties with CTO crossing and comorbidities (left ventricular disfunction, particularly in the donor vessel is being instrumented during retrograde procedures).^1, 13-15^ How can we identify patients who are more likely to need MCS? We recently developed risk scores for estimating the risk of periprocedural in-hospital major adverse cardiovascular events (MACE), mortality, pericardiocentesis, and acute myocardial infarction (MI) in patients undergoing CTO PCI ^11^, but there are currently no risk scores for assessing the need for urgent MCS in CTO PCI.

There is limited data regarding MCS in CTO PCI. In a prior publication of the PROGRESS CTO registry, MCS was used electively in 69 procedures (4%) and urgently in 22 procedures (1%).^16^ In a retrospective cohort, elective MCS support with the Impella 2.5 or CP was used in 57 CTO PCIs (2%). Technical (87.7%) and procedural (75.4%) success were high, but so was the risk of periprocedural complications occurred: vascular injury (5.3%), all-cause death (5.3%), major bleeding (3.5%), stroke (1.8%), and coronary perforation resulting in tamponade (1.8%).^17^ An analysis of the National Inpatient Sample (NIS) between 2008-2014 found that MCS was utilized in 2% of hospitalizations with CTO-PCI (n=93,109). MCS utilization, both elective and urgent, increased during the study period. While overall in-hospital mortality was 2%, it was 25.9% among MCS patients compared with 1.6% in patients who did not need MCS (p<0.0001). Patients requiring MCS have more comorbidities and more likely to be in cardiogenic shock, limiting their tolerance of procedural complications. An additional explanation could be the development of acute kidney injury during MCS hospitalizations, which was higher in patients who received MCS.^18^ Azzalini et al. evaluated the early and one-year outcomes of 250 Impella-supported (Impella 2.5 or CP) high-risk nonemergent PCIs in a single-center retrospective study (15% of the lesion were CTOs). After propensity matching the incidence of MACE was higher in the MCS group (26.8% vs. 13.2%, p<0.001), as was the incidence of periprocedural MI (14.0% vs. 6.4%, p= 0.005), major bleeding (6.8% vs. 2.8%, p=0.04), and need for blood transfusion (11.2% vs. 4.8%, p= 0.008). In addition, in-hospital mortality trended numerically higher in the Impella-supported group (3.2% vs. 1.2%, p= 0.13). There were no differences in the incidence of MACE (31.2% vs. 27.4%, p=0.78) or any of its individual components between Impella-supported patients and controls at one-year follow-up.^19^ A retrospective, observational, multi-center (10 hospital) registry included 157 patients who underwent high risk PCI with Impella support (14% CTOs). During 180-day follow-up, MACE occurred in 34 patients (23%), 27 patients (18%) died, 9 patients (6%) sustained a ST-elevation myocardial infarction, and 4 patients (3%) suffered a stroke.^20^

In a single-center study of 13 prophylactic Tandem Heart-supported CTO PCIs the most common reason for hemodynamic support was use of the retrograde approach in the setting of left ventricular dysfunction (38%). Technical success was high (92%) despite high lesion complexity. Procedural success was 77%, there were no major bleeding complications, but one patient developed an arteriovenous fistula at the arterial cannula insertion site, one patient had a coronary perforation with hemodynamic compromise requiring pericardiocentesis, and one patient died of cardiogenic shock, secondary to right ventricular wall hematoma.^21^

Use of the PROGRESS CTO urgent MCS score may facilitate patient selection for prophylactic hemodynamic support optimizing the risk benefit ratio of the procedure.

### Study limitations

The primary limitation of this study is the relatively small number of procedures requiring urgent MCS. These are rare events and given limited available data, we tackled this issue using a bootstrap resampling to develop the least complex-most informative model. Furthermore, the out-of-sample validation was limited to CTO PCIs not requiring urgent MCS, so only model specificity could be estimated. Other limitations of our study include its observational design, lack of clinical event adjudication and core laboratory analyses, and using data from high-volume, experienced PCI centers with a record of performing urgent MCS, which limits the generalizability of our findings. The score performance will need to be re-evaluated as more data become available.

## CONCLUSION

In conclusion, use of the PROGRESS CTO urgent MCS score may facilitate patient selection for prophylactic MCS and optimize the risk benefit ratio of CTO PCI.

## Data Availability

Data not available.

## Acknowledgments

Study data were collected and managed using Research Electronic Data Capture (REDCap) electronic data capture tools hosted at the Minneapolis Heart Institute Foundation (MHIF), Minneapolis, Minnesota. REDCap is a secure, web-based application designed to support data capture for research studies, providing: (1) an intuitive interface for validated data entry; (2) audit trails for tracking data manipulation and export procedures; (3) automated export procedures for seamless data downloads to common statistical packages; and (4) procedures for importing data from external sources.

---

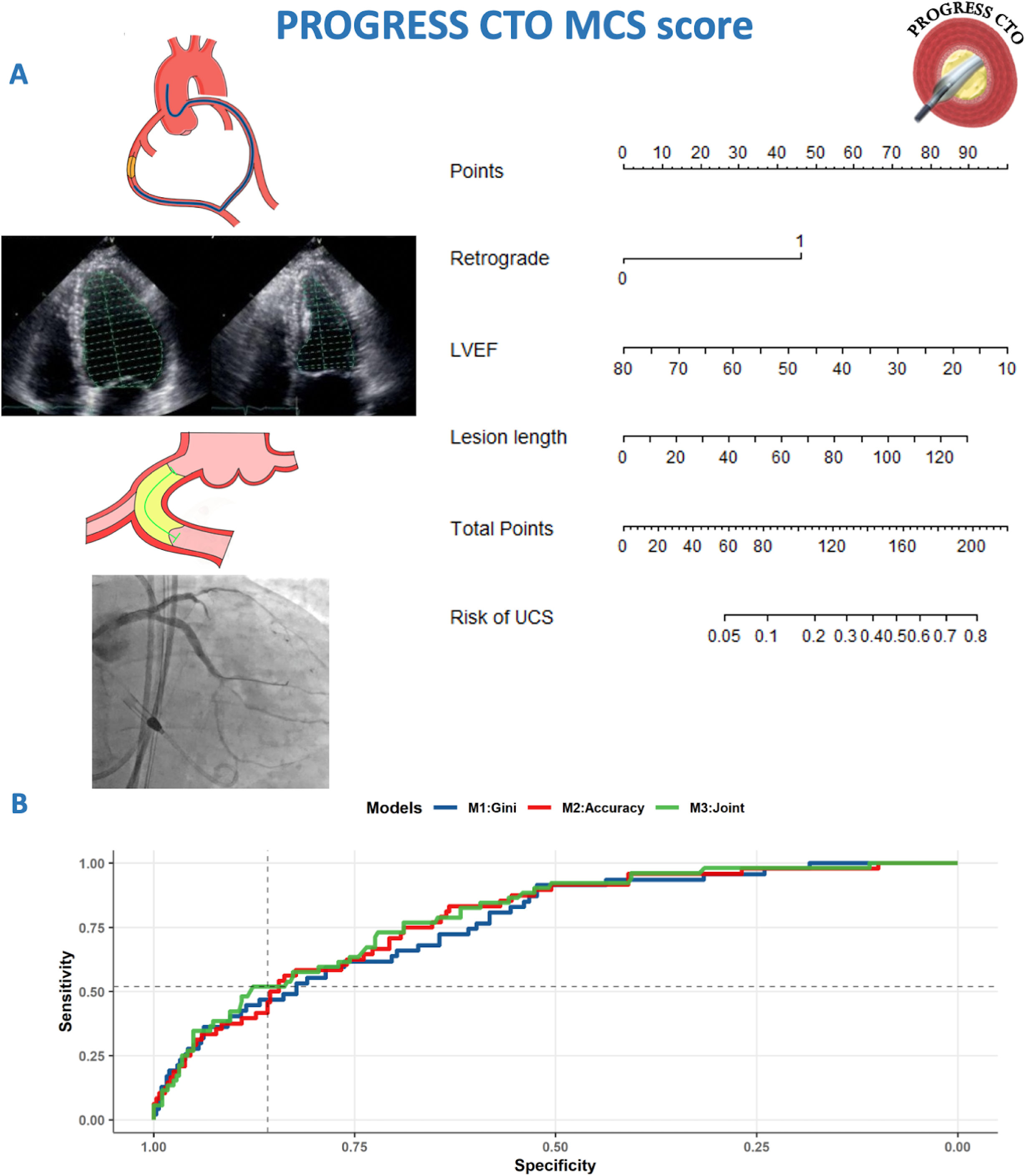

